# IgG2 Galactosylation is related to higher antibody dependent enhancement for dengue in cross-reactive antibodies from Sars-CoV-2

**DOI:** 10.64898/2026.06.22.26356250

**Authors:** Sebastian Reinig, Chin Kuo, Shin-Ru Shih

**Author notes:** Corresponding author is Sebastian Reinig.

## Abstract

Cross-reactive antibodies against dengue virus are known to cause antibody-dependent enhancement (ADE) of infection or disease severity under specific conditions. In our previous study, we showed that primary immunization with the COVID-19 vaccine induces induces cross-reactive IgG causing ADE against dengue. In the present study, we investigated the influence of IgG Fc-glycosylation (analyzed by LC-MS/MS) on ADE mediated by cross-reactive IgG against dengue from IgG against SARS-CoV-2. We found a clear correlation between anti-DENV2 E IgG2 galactosylation and the ADE capacity of cross-reactive IgG against dengue in individuals vaccinated against COVID-19. IgG2 sialylation increased over time; however, it was not correlated with ADE capacity. This phenomenon was restricted to IgG2, whereas anti-DENV2 E IgG1 Fc-glycosylation remained stable after COVID-19 vaccination.

## Introduction

Dengue disease poses a major public health threat, with an increasing incidence of over 60 million cases reported worldwide annually and more than 36,000 deaths^1,2^. Dengue virus is a flavivirus and is transmitted by mosquitos. While most infections are asymptomatic or cause mild disease, a small proportion of cases can progress to severe dengue (dengue hemorrhagic fever), which may require hospitalization and can be fatal^3^. Dengue virus exhibits strong cross-reactivity with antibodies induced by other dengue serotypes, related flaviviruses, and even unrelated viruses such as poliovirus or SARS-CoV-2^4–7^. Cross-reactive IgG antibodies against dengue are typically non-neutralizing. These antibodies can facilitate viral entry via Fcγ receptors instead of providing protection, thereby enhancing viral replication. This phenomenon is known as antibody-dependent enhancement (ADE)^8^. In our previous study, we observed that primary immunization with the COVID-19 vaccine induces strong antibody-dependent enhancement (ADE) mediated by IgG after the first dose, an effect that declines with each subsequent dose^6^. We have not identified any factor that correlates with the observed differences in ADE capacity. Each IgG antibody has two conserved N-glycosylation sites in the Fc domain, which can modulate Fcγ receptor affinity. This is particularly important for ADE of dengue, and prior studies have shown that Fc-fucosylation can increase ADE, thereby enhancing infection and disease severity^8–10^. We therefore investigated the Fc-glycosylation of cross-reactive anti-dengue IgG antibodies from COVID-19-vaccinated individuals in our previous study.

## Methods

### Sample

In total, 63 serum or plasma samples were collected from individuals before vaccination and after the first, second, and third doses of the following vaccines: the adenoviral vector vaccine ChAdOx1 nCoV-19 (AstraZeneca, Cambridge, UK); the protein subunit vaccine MVC-COV1901 (Medigen, Frederick, MD, USA); the mRNA vaccines mRNA-1273 (Moderna, Norwood, MA, USA) and BNT162b2 (BioNTech/Pfizer, Mainz, Germany); and Omicron-era vaccines targeting XBB and BA.4.5 (**Table 1**). Samples from Taiwan were collected from individuals at Chang Gung Memorial Hospital and Chang Gung University (ethics approval number: 202001041A3C).

**Table 1.**
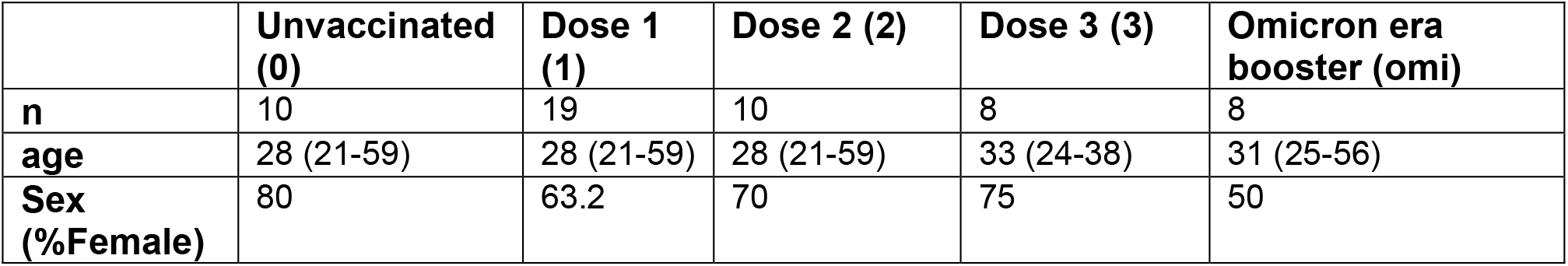
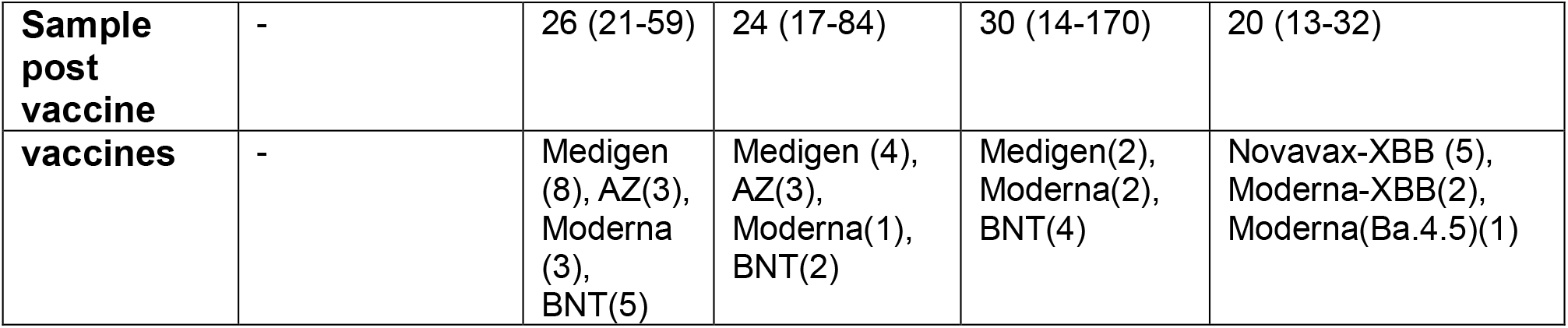
Sample collection for each group, reported as median, minimum, and maximum values. Individuals received ChAdOx1 nCoV-19 (AstraZeneca, AZ), MVC-COV1901 (Medigen), or the mRNA vaccines mRNA-1273 (Moderna) or BNT162b2 (BNT),

### Antibody isotype enzyme-linked immunosorbent assay

The data presented originated from our prior study^6^. Envelope protein from DENV serotype 2 (Sino Biological, Beijing, China) was immobilized at 0.2 mg/mL per well in PBS on a 96-well flat-bottom polystyrene microplate overnight. A blocking solution of 2.5% bovine serum albumin (BSA) in PBST (PBS with 0.05% Tween 20) was applied. All samples were diluted in PBST under identical conditions: 1:200 for anti-E IgG subclasses. Horseradish peroxidase (HRP)-conjugated anti-human secondary antibodies were used at a 1:5000 dilution in 2.5% BSA in PBST: IgG1, IgG2, IgG3, and IgG4 (SouthernBiotech, Birmingham, AL, USA); IgM (Sigma-Aldrich, St. Louis, MO, USA)

### Anti-DENV2 envelope IgG isolation

BcMagTM Tosyl Activated Magnetic Beads (BioClone) were used to purify antigen-specific IgG (DENV envelope protein serotype 2) from the serum. For coupling of the E protein, 100 μg of E protein (Sino Biological, Beijing China) was bound to 30 mg of Tosyl magnetic beads. The beads were then divided equally for each sample, with coupling beads containing 2.5 μg of DENV2 E protein used for the serum. Antibodies were eluted with 0.1% trifluoroacetic acid. The DENV2 E protein-specific IgGs were subsequently analyzed using mass spectrometry.

### Analysis of the Fc-glycosylation by Mass spectrometry

LC-MS/MS analysis was performed as described in our prior study. Here, Anti-E IgG was used instead. Protein concentration was determined by measuring 5 μg of protein on a Nanodrop spectrophotometer at 280 nm. For each glycoform, the signal intensities of the individual modifications (fucosylation, galactosylation, sialylation, and bisection) were summed. This sum was then divided by the total signal intensity of all Fc-N297 glycoforms (used as the denominator) to calculate relative abundances.

### ADE assay for serum antibodies

The data were obtained from our prior study^6^. DENV type 2 was incubated at a multiplicity of infection (MOI) of 0.001 with serum diluted 1:1000 for 30 min on ice. The mixture was then incubated with 10^5^ THP-1 cells for 90 min. The THP-1 cells were washed twice, and the supernatant was collected after 48 h for quantitative PCR (qPCR).

### qPCR of Dengue 2 virus

The data for the samples from this experiment was obtained from our former study^6^ and the details for the method can be found in. For the qPCR analysis, the ΔΔCt method was applied, with gene expression fold changes expressed as log_2_ values.

### Statistics

Statistical analyses were performed using R software (ver. 4.3.1) and the dplyr package. All figures were created using R. For violin plots, the median is given. The Mann–Whitney U test was used for comparisons between two samples. The pairwise Wilcoxon test was used to compare multiple groups. Values of p < 0.05 were considered significant. Pearson correlation coefficients were measured using the cor.test function in R to evaluate relationships between IgG profile parameters and other variables (IgG subclass glycoforms, and IgG subclass titer).

## Results

### IgG2 Galactosylation is specifically correlated to antibody-dependent enhancement of cross-reactive antibodies from COVID-19 vaccination

The Fc-N297 glycosylation profile of anti-DENV2 Envelope IgG1 and IgG2 was measured in 62 samples from individuals who were either unvaccinated or had received 1–3 doses of the Wuhan-variant COVID-19 vaccine at the time of sample collection, as well as from individuals vaccinated with Omicron-era vaccines (**Table 1, Figure 1A, B**). Fucosylation was significantly lower on anti-DENV2 E IgG1 compared with IgG2 (92% vs. 99%, respectively), whereas galactosylation was significantly higher on IgG1 than on IgG2 (77% vs. 37.3%, respectively). Bisection and sialylation levels did not differ significantly between IgG1 and IgG2 (bisection: 3.7% vs. 2.7%; sialylation: 2.6% vs. 7.9%). No significant differences in Fc-glycosylation were observed for anti-DENV2 E IgG1 across the different vaccination groups (**Figure 1A**). In contrast, anti-DENV2 E IgG2 showed a significant increase in galactosylation from the unvaccinated cohort (15.6%) to the first dose (53.8%), followed by a decrease after the second (29.1%) and third doses (21.8%). Samples from individuals who received the XBB booster exhibited the highest galactosylation (65.3%) (**Figure 1B**). Sialylation on IgG2 also increased with the number of doses (1.2%, 11.4%, and 36% after the first, second, and third doses, respectively; **Figure 1B3**). IgG subclass titers did not differ significantly between cohorts, except for IgG4, which was significantly elevated after the third dose (**Figure 1C**). Antibody-dependent enhancement (ADE) capacity, measured as the fold change in viral replication relative to virus incubation with THP-1 monocytes alone, was significantly increased after the first dose, declined after the second and third doses, and rose again in individuals who received the Omicron booster (**Figure 1D**). Only galactosylation showed a significant correlation with ADE capacity (r^2^ = 0.26, p < 0.01; **Figure 1E**).

**Figure 1.**
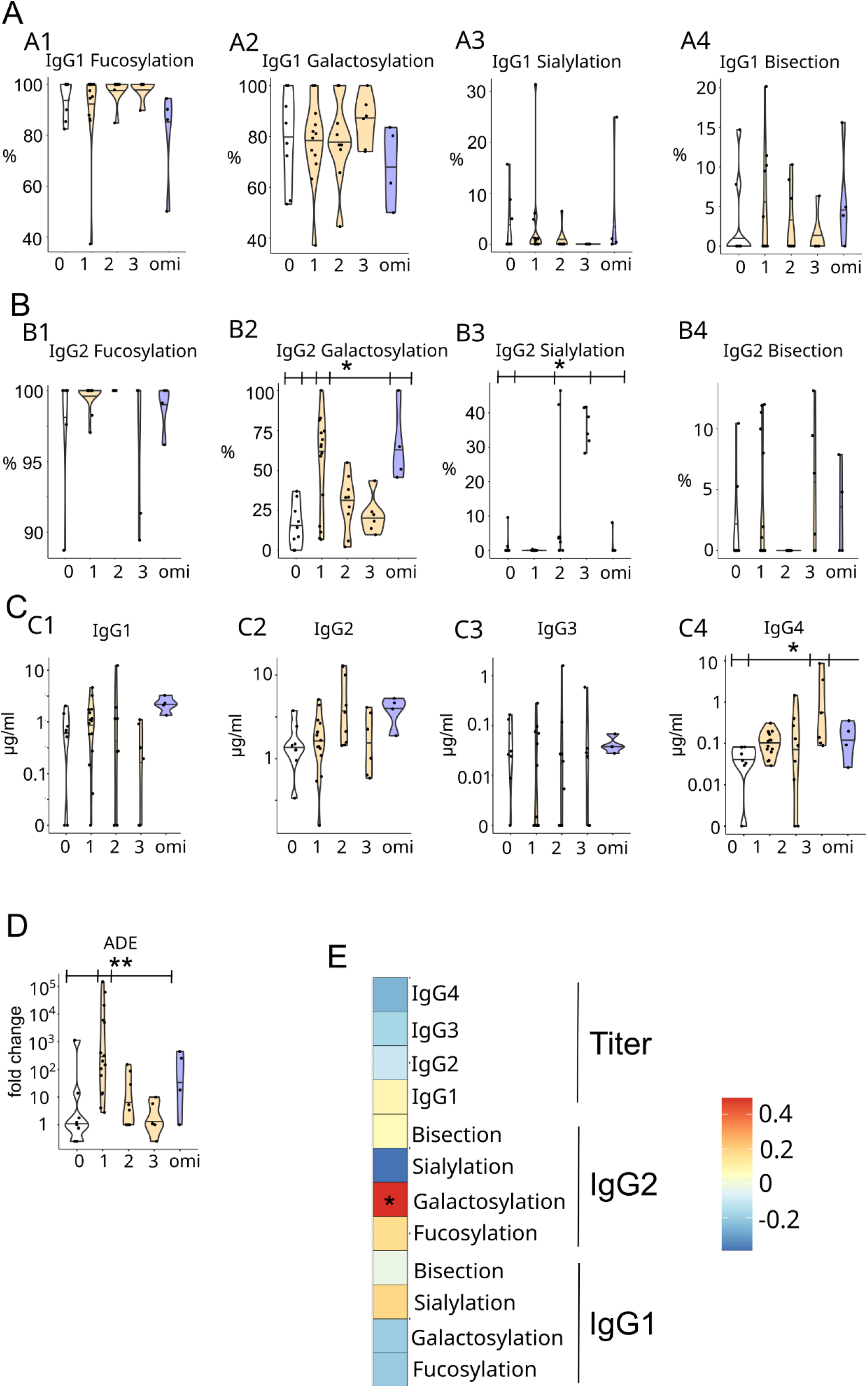
Fc-N297 glycosylation modifications of anti-DENV2 Envelope (E) IgG1 (**A**) and IgG2 (**B**). Shown are the relative levels of fucosylation, galactosylation, sialylation, and bisection. (**C**) Anti-DENV2 E IgG subclass titers. (**D**) Antibody-dependent enhancement (ADE) capacity, expressed as the fold change in DENV2 viral RNA levels in THP-1 cells treated with serum/plasma samples relative to THP-1 cells infected with DENV2 virus alone. (**E**) Pearson correlation coefficient of the glycosylation and subclass titer of anti-DENV2 to log-normalized ADE capacity. Statistical significance was determined by Kruskal–Wallis test (*p < 0.05, **p < 0.01). 0: unvaccinated; 1–3: 1st to 3rd dose of Wuhan-variant COVID-19 vaccine; Omi: Omicron-era booster.

## Discussion

Our study is among the first to demonstrate a functional role of IgG2 galactosylation in vitro and provides one of the first indications of a broader biological effect of IgG2 glycosylation in general. Research on the influence of IgG glycosylation in dengue and, more broadly, in infectious diseases has focused almost exclusively on IgG1^10,12,13^. The potential mechanism by which galactosylation could influence ADE of anti-DENV2 E IgG2 is unknown. Fc-galactosylation is known to increase Fc receptor affinity and enhance processes such as ADCP, which is a mechanism by which dengue virus can exploit antibodies for ADE ^13,14^. However, our previous study found no correlation between Fcγ receptor affinity and ADE capacity. We also found that ADE mediated by cross-reactive IgG is influenced by the complement system^6^.IgG2 exhibits low affinity for C1q, the first component of the classical complement cascade. Consistent with this, a prior study found no influence of complement on the ADE capacity of anti-DENV2 IgG2 ^15^. Although this is only a correlative finding, additional experiments with samples from COVID-19-vaccinated individuals — using IgG2-depleted or purified preparations, as well as glycoengineered cross-reactive anti-dengue IgG2 — are needed to confirm that galactosylation is a contributing factor. Furthermore, the underlying mechanism and the epitopes targeted by these cross-reactive IgG antibodies are currently unknown. This preliminary publication is based on a limited number of unpaired samples. A larger study with paired samples is therefore needed to confirm these findings.

## Data Availability

All data produced in the present study are available upon reasonable request to the authors

